# Added value of cell-free DNA over clinical and ultrasound information for diagnosing ovarian cancer

**DOI:** 10.1101/2024.06.20.24309215

**Authors:** Adriaan Vanderstichele, Jolien Ceusters, Daniela Fischerova, Antonia Testa, Wouter Froyman, Chiara Landolfo, Ruben Heremans, Francesca Moro, Anne-Sophie Van Rompuy, Thaïs Baert, Els Van Nieuwenhuysen, Toon Van Gorp, Ignace Vergote, Pieter Busschaert, Tom Venken, Diether Lambrechts, Joris Robert Vermeesch, Tom Bourne, An Coosemans, Ben Van Calster, Dirk Timmerman

## Abstract

**Background:** We previously proposed two cfDNA-based scores (genome-wide z-score and nucleosome score) as candidate non-invasive biomarkers to further improve pre-surgical diagnosis of ovarian malignancy. We aimed to investigate the added value of these cfDNA-based scores to the predictors of the ADNEX model (Assessment of Different NEoplasias in the adnexa) to estimate the risk of ovarian malignancy.

**Methods:** 526 patients with an adnexal mass scheduled for surgery were consecutively recruited in three oncology referral centers. cfDNA-based scores were calculated in pre-operative plasma samples. Logistic regression models were fitted for ADNEX predictors alone and after adding cfDNA scores. We reported likelihood ratio tests, the area under the Receiver Operating Characteristic curve (AUC), sensitivity, specificity, and Net Benefit for thresholds between 5% and 40%.

**Results:** The study included 272 benign, 86 borderline, 36 stage I invasive, 113 stage II-IV invasive, and 19 secondary metastatic tumors. The likelihood ratio tests for adding the cfDNA variables to the ADNEX model were statistically significant (p<0.001 for ADNEX without CA125, p=0.001 for ADNEX with CA125). The accompanying increases in AUC were 0.013 and 0.003. Net Benefit, sensitivity and specificity were similar for all models. The increase in Net Benefit at the recommended 10% threshold estimated risk of malignancy was 0.0017 and 0.0020, respectively. According to these results, adding cfDNA markers required at least 453 patients per additional true positive.

**Conclusion:** Although statistically significant, the addition of the cfDNA scores to the ADNEX model do not improve the ADNEX model in a clinically meaningful way.

## INTRODUCTION

Adnexal masses are common, with some studies reporting a lifetime risk of 5-10% for women to undergo surgery for a suspected ovarian malignancy(1). During follow-up of these adnexal masses, gynecologists are confronted with a diagnostic dilemma, as they need to balance the disadvantage of undergoing surgery (i.e., risk of complications, loss of fertility and health-economic considerations) against the risk of missing the diagnosis of an invasive tumor.

The ADNEX (Assessment of Different NEoplasias in the adneXa) risk model was developed by the International Ovarian Tumor Analysis (IOTA) group as a multiple outcome prediction method to estimate the probability that an adnexal mass is benign, borderline, stage I invasive ovarian cancer, stage II-IV invasive ovarian cancer, or secondary metastatic cancer based on clinical and ultrasound data(2). This model has been validated extensively, indicating that it discriminates between benign and malignant masses with areas under the receiver operating characteristic curve (AUC) of 0.93-0.94(3,4). ADNEX currently represents a clinical standard to predict ovarian malignancy.

Current developments in early cancer detection increasingly focus on cell-free DNA (cfDNA), usually obtained from plasma samples. Previously, the presymptomatic detection of cancers was observed in women undergoing non-invasive prenatal testing(5).

Low concentrations of cfDNA are present in plasma of healthy individuals as short double-stranded DNA fragments; 70–90% of this cfDNA is derived from leukocytes, while the remaining amounts originate from several other organs, such as the liver(6,7). In patients with (invasive) cancer, a highly variable percentage of cfDNA originates from the tumor. Efforts to characterize this tumor-specific fraction (ctDNA, i.e. circulating tumor DNA) focus on the detection of tumor-specific genetic variation, i.e. somatic mutations and copy number alterations (CNAs), as well as epigenetic features of cfDNA such as tumor-specific patterns of DNA methylation and specific patterns of cfDNA fragmentation caused by (tumor) cell-type specific patterns of nucleosome positioning(6–12).

In this context, we previously proposed two cfDNA-based biomarkers, the genome-wide z-score(13) and the nucleosome score(14) as candidate non-invasive biomarkers to further improve pre-surgical detection of ovarian malignancy. These two biomarkers are calculated as two separate read-outs of low-coverage whole genome sequencing (LC-WGS) on plasma-derived cfDNA and results indicated that these biomarkers might provide additional diagnostic information. While genome-wide z-score analysis aims to detect tumor-specific chromosomal instability in cfDNA, the quantification of nucleosome footprinting of cfDNA using the nucleosome score aims to detect cancers independently from the presence of chromosomal instability (e.g. subset of non-high grade serous invasive ovarian cancer).

It is hypothesized that cfDNA-based biomarkers may display increased test specificity as they inherently include tumor-specific features. Therefore, our aim was to evaluate the added diagnostic value of these cfDNA-based biomarkers in combination with established clinical and ultrasound predictors incorporated into the ADNEX model.

## METHODS

### Study design and participants

This prospective cohort study consecutively enrolled patients who presented with an adnexal (ovarian, paraovarian, tubal, paratubal) mass at three centers: University Hospitals Leuven, Leuven, Belgium (recruitment 6/2015-9/2019); Charles University, Prague, Czech Republic (recruitment 12/2017-9/2019); Università Cattolica del Sacro Cuore, Rome, Italy (recruitment 1/2019-7/2019). All patients underwent a standardized ultrasound examination following the IOTA methodology(15), and were scheduled for surgery by the treating physician. Exclusion criteria were (1) surgery more than 120 days after ultrasound examination, (2) age <18 years at the ultrasound examination, (3) ongoing pregnancy, (4) known simultaneous and/or previous malignancies in the previous five years, (5) surgery for the suspected mass elsewhere prior to inclusion, (6) refusal of preoperative transvaginal ultrasonography or blood withdrawal, and (7) denial or withdrawal of informed consent. This study was part of the transIOTA study, and excludes earlier transIOTA data that were used in the derivation of the cfDNA biomarkers(13,14). TransIOTA was an amendment study to the large and non-overlapping IOTA5 and IOTA7 studies. Approval for the transIOTA study was obtained from the Research Ethics Committee of the University Hospitals KU Leuven (Leuven, Belgium) as coordinating centre (S51375/NCT01698632 and S59207/NCT02847832) and the local ethics committee of each contributing center. All patients gave written informed consent.

This study was reported according to the Reporting recommendations for tumor MARKer prognostic studies (REMARK) checklist(16).

### Ultrasound examination

Transvaginal ultrasound examination was performed according to the standardised research protocol, using the IOTA terms and definitions(15).

All ultrasound examiners were experienced and collected the variables used in the ADNEX model(2). The model includes the age of the patient, the type of center (oncology referral unit vs other unit), six ultrasound variables (maximal diameter lesion, maximal diameter of largest solid part, more than 10 cyst locules, number of papillations, acoustic shadows, ascites), and the serum CA125 level as an optional predictor. This information was collected using Clinical Data Miner, a secure web-based data collection tool(17).

### Biological sample processing and bio-informatics

A blood sample (two Cell-Free DNA BCT® tubes from Streck, Inc., ref 218997) was taken at the time of the ultrasound examination or the day prior to surgery. Plasma was prepared and cfDNA was extracted in two separate labs (Laboratory of Translational Genetics (VIB-KU Leuven) and Laboratory for Cytogenetics and Genome Research (LCGR-KU Leuven)), using similar and standardized protocols as previously described(13,18). Quality control included analysis of similar samples by both labs. Briefly, DNA single-end sequencing libraries were prepared and all samples were subjected to low-coverage short-read whole-genome sequencing, with a median read count of 9.3·10^6^ reads per sample. Raw sequencing reads were mapped to the human reference genome Hg19 using BWA v0.7.1(19). Duplicate and low-quality reads were removed by Picard Tools v1.11 and Samtools v0.1.18, respectively(20). 170/526 samples were processed using the VIB protocol, 296 using the LCGR protocol, and 60 using both protocols (the analysis used results from LCGR).

The *genome-wide z-score* assesses chromosomal instability(13). The genome was divided in 1000 kbp bins, excluding sex chromosomes. Reads were counted in each bin and adjusted for the total number of reads, GC-content and mappability. The bin values were smoothened by taking moving window averages of 50 adjacent bins. Then *z*-scores were calculated for each window using the distribution of 44 healthy female individuals as a reference. Subsequently, a single genome-wide *z*-score was calculated for each sample as the *z*-score (again using healthy individuals as a reference) of the sum of squares of all window *z*-values.

The *nucleosome score* quantifies genome-wide variation of nucleosome footprints(14). The start positions of 51 bp reads—representing the boundaries of circulating cfDNA fragments— were compared to a map of nucleosome positions found in plasma of 125 healthy female individuals(21). Distances on autosomes between each read start and the nearest nucleosome center from the reference list, within a [-300, +300] bp range, were recorded. Using a multinomial stochastic model, trained on plasma samples of 125 healthy individuals and 43 patients with (high-grade serous) ovarian cancer, as a reference, we could calculate a nucleosome score for each plasma sample. An estimated nucleosome score near 0 should correspond better to the typical sample of a healthy individual, whereas an estimated nucleosome score near 1 should correspond better to a sample of a patient with ovarian cancer.

### Statistical analysis

A prespecified statistical analysis plan was followed. The genome-wide z-score, nucleosome score and CA125 each had 3 to 4% missing values. We assumed that these occurred completely at random due to technical issues (PCR problems, low number of reads, insufficient plasma or serum quantity). We imputed missing values (**Supplementary Appendix 1**).

Correlations between cfDNA scores and ADNEX predictors were calculated. Plasma samples were analysed using one of two protocols, with 60 being analysed using both protocols. For these samples, we assessed the agreement between protocols using Bland-Altman analysis.

The univariable analysis of the cfDNA scores involved AUCs with 95% confidence intervals (CI)(22). The multivariable analysis involved logistic regression models using Firth’s correction to assess the added value of the cfDNA scores above established clinical and ultrasound predictors included in the ADNEX model: age, maximum diameter of lesion, proportion of solid tissue, more than 10 cyst locules, number of papillary projections, acoustic shadows, ascites, and CA125 (**Supplementary Appendix 1**). The cfDNA scores were modelled using restricted cubic splines with three knots to allow for nonlinear associations(23). Because CA125 is an optional variable in ADNEX, we fitted different models with the following set of predictors:

1. ADNEX predictors without CA125 (model 1)
2. Model 1 predictors + genome-wide z-score (model 2)
3. Model 1 predictors + nucleosome score (model 3)
4. Model 1 predictors + genome-wide z-score and nucleosome score (model 4)
5. ADNEX predictors including CA125 (model 5)
6. Model 5 predictors + genome-wide z-score (model 6)
7. Model 5 predictors + nucleosome score (model 7)
8. Model 5 predictors + genome-wide z-score and nucleosome score (model 8)

We evaluated the added value of cfDNA biomarkers first using likelihood ratio tests for model 4 vs 1 and model 8 vs 1. Then, we assessed model performance through AUC, sensitivity, specificity and Net Benefit. We used the enhanced bootstrap (1000 bootstraps) to correct performance measures for optimism(23,24) We used the DeLong method(25) to calculate CIs on the difference in AUC. Sensitivity and specificity were calculated for prespecified thresholds 1%, 5%, 10%, 20%, 30%, 40% and 50%. Net Benefit was calculated for thresholds between 5% and 40% to assess clinical utility to decide between specialized vs conservative surgery(26). The inverse of the difference in Net Benefit is the ‘Test Tradeoff’, the number of patients per extra true positive (patient with a malignancy for which the model suggested specialized surgery)(27,28).

Based on simulations, we assumed that our sample size had ≥80% power for the likelihood ratio tests when adding cfDNA variables increased the AUC by ≥0.006 (**Supplementary Appendix 1**). The sample size was also sufficient for the multivariable models (**Supplementary Appendix 1**). All statistical analyses were performed in R version 4.1.0(29), using packages mice, auRoc, logistf and rms.

## RESULTS

### Study dataset

Between June 2015 and September 2019, 573 patients were enrolled from three contributing centers (**Figure 1**). 47 patients met an exclusion criterion, resulting in 526 patients (334 from Leuven, 102 from Prague and 90 from Rome). 272 patients had a benign adnexal mass, and 254 patients had a malignant adnexal tumor (86 (16%) borderline carcinoma, 149 (28%) primary invasive ovarian cancer and 19 (4%) secondary metastases to the ovary from other primary origins)(**table 1**). Median patient age was 55 years. 61% of patients were postmenopausal.

**Figure 1.**
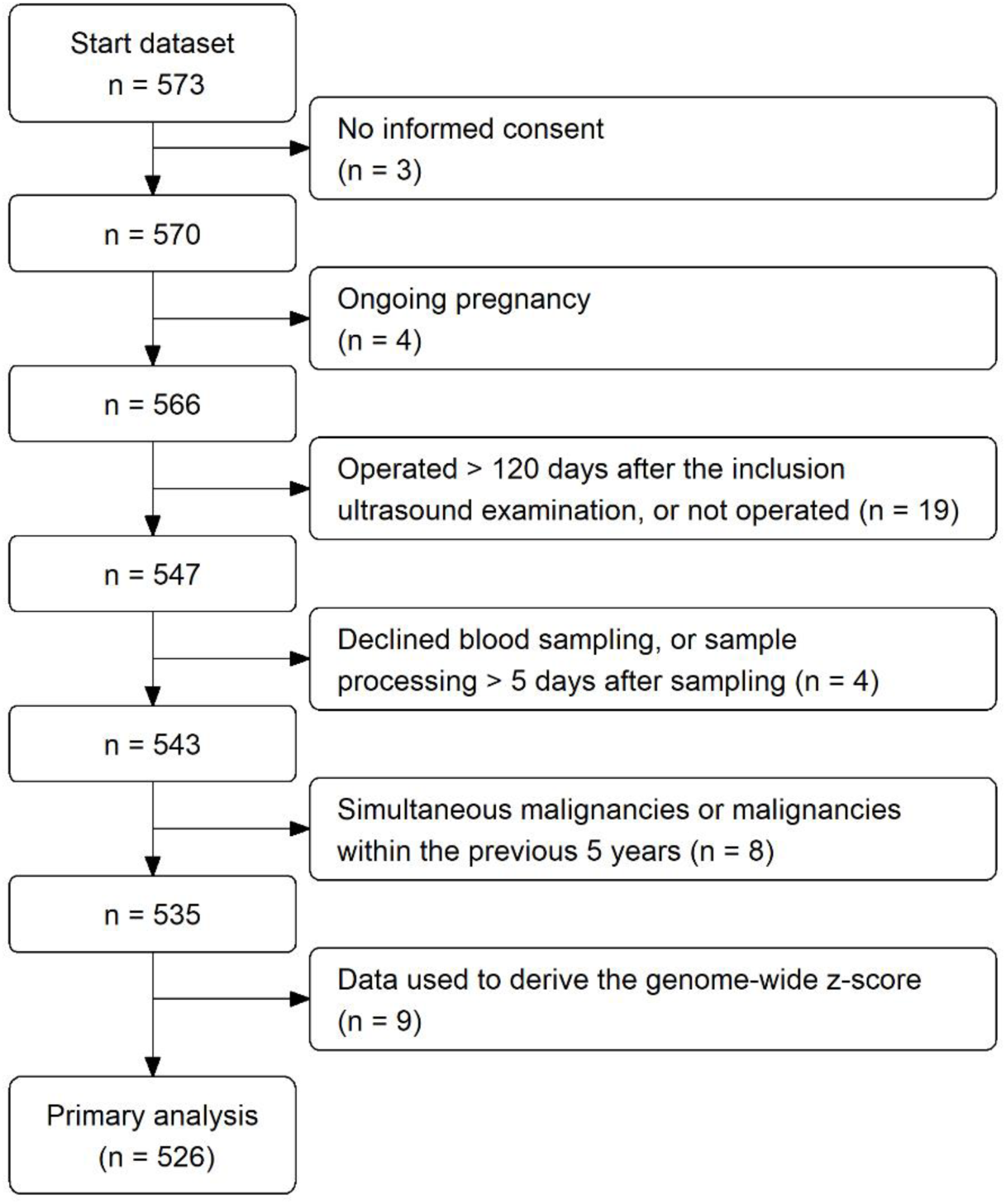
Flowchart of the patients included in the analysis.

**Table 1.** Baseline characteristics (n = 526). Patient, histopathological and ultrasound variables. IQR = interquartile range.

| **Variable** | **Median (IQR) (range), or n (%)** |
| --- | --- |
| Patient age at recruitment (years) | 55 (44–65) (19–88) |
| Postmenopausal | 320 (61%) |
| Maximum diameter of lesion (mm) | 73 (48–114) (7–459) |
| Presence of solid components | 364 (69%) |
| Maximum diameter of largest solid component (mm)<sup>a</sup> | 43 (24–70) (3–230) |
| Presence of > 10 cyst locules | 86 (16%) |
| Number of papillary projections |  |
| 0 | 372 (71%) |
| 1 | 72 (14%) |
| 2 | 19 (4%) |
| 3 | 10 (2%) |
| At least 4 papillations | 53 (10%) |
| Presence of acoustic shadows | 204 (39%) |
| Presence of ascites | 79 (15%) |
| Presence of metastasis | 70 (13%) |
| Bilateral masses | 148 (28%) |
| Serum CA125 (kU/L)<sup>b</sup> | 28 (13–153) (3–24137) |
| Genome-wide z-score<sup>b</sup> | 0.66 (-0.02–2.35) (-0.93–4031) |
| Nucleosome score<sup>b</sup> | 0.11 (0.002–0.36) (0.0002–1) |
| Ultrasound examiner's subjective impression |  |
| Probably benign | 130 (25%) |
| Certainly benign | 49 (9%) |
| Probably malignant | 100 (19%) |
| Certainly malignant | 160 (30%) |
| Uncertain | 87 (17%) |
| Outcome |  |
| Benign | 272 (52%) |
| Borderline | 86 (16%) |
| Stage I invasive ovarian cancer | 36 (7%) |
| Stage II – IV invasive ovarian cancer | 113 (21%) |
| Secondary metastatic cancer | 19 (4%) |

<sup>a</sup> Only for tumors with a solid component
<sup>b</sup> There were 18 (3%) missing values for the genome-wide z-score and the nucleosome score, and 20 (4%) missing values for CA125.

### Cell-free DNA (cfDNA)-based biomarkers

We observed low values for nucleosome and genome-wide *z*-scores in patients with benign disease (**supplementary table 1**, **supplementary figures 1-2**). Both cfDNA-based scores for patients with borderline carcinomas were similar to those from patients with benign disease. Patients with stage II-IV invasive disease displayed a high median genome-wide z-score of 7.57 and a high median nucleosome score of 0.44. Patients with metastatic disease had low to moderately elevated median cfDNA scores, comparable to patients with stage I invasive disease. The Spearman correlation between the 2 cfDNA-based scores was 0.40. Correlations between the ADNEX variables and the cfDNA scores are shown in **supplementary table 2**.

**Figure 2.**
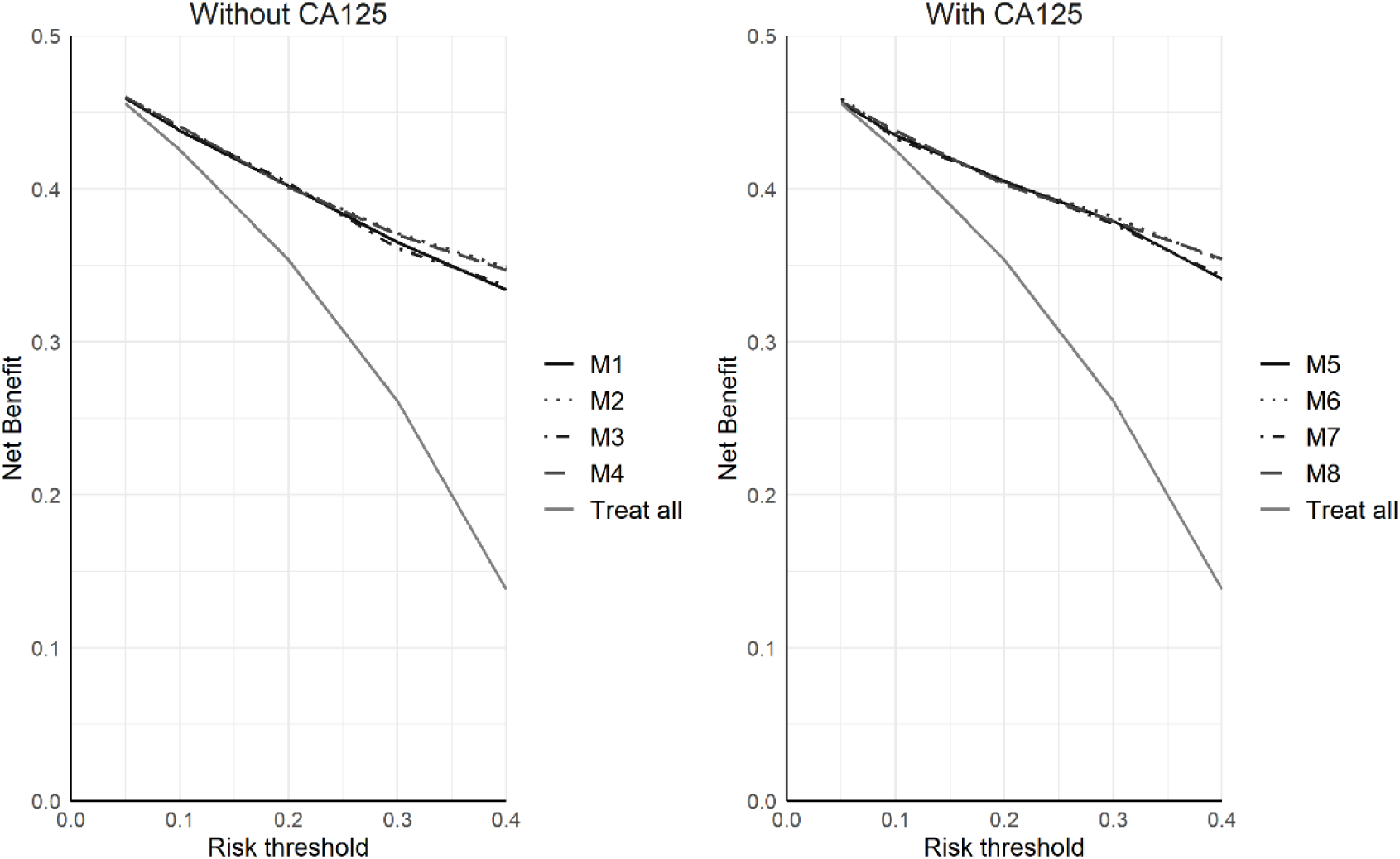
Decision curves of the different models for the comparison benign (n=272) versus malignant (n=254).

Based on Bland-Altman analysis, the average difference for genome-wide z-score and nucleosome score was -29.39 (95%CI: -72.02; 13.23) and 0.002 (95%CI: -0.016; 0.019)(**supplementary figures 3-4**).

### Performance of cfDNA-based prediction of malignancy

The AUC to distinguish benign from malignant (including borderline, invasive carcinoma, and ovarian metastasis) was 0.73 (95% CI: 0.68–0.77) for the genome-wide z-score and 0.64 (95% CI: 0.59–0.69) for the nucleosome score. The AUC values for specific comparisons are shown in **table 2**.

**Table 2.** Areas under the receiver operating curve (AUCs) with 95% confidence intervals for the discrimination between subtypes of tumors based on the cell-free DNA (cfDNA) biomarkers.

| **Subtype comparison** | **Genome-wide z-score** | **Nucleosome score** |
| --- | --- | --- |
| Benign vs borderline | 0.59 [0.52; 0.65] | 0.53 [0.46; 0.60] |
| Benign vs stage I | 0.66 [0.56; 0.75] | 0.55 [0.45; 0.65] |
| Benign vs stage II-IV | 0.86 [0.82; 0.90] | 0.75 [0.69; 0.80] |
| Benign vs metastatic | 0.66 [0.52; 0.77] | 0.64 [0.51; 0.76] |
| Borderline vs stage I | 0.58 [0.47; 0.68] | 0.53 [0.41; 0.63] |
| Borderline vs stage II-IV | 0.82 [0.76; 0.87] | 0.73 [0.65; 0.79] |
| Borderline vs metastatic | 0.60 [0.45; 0.73] | 0.63 [0.48; 0.75] |
| Stage I vs stage II-IV | 0.78 [0.68; 0.85] | 0.70 [0.59; 0.78] |
| Stage I vs metastatic | 0.54 [0.38; 0.69] | 0.59 [0.43; 0.74] |
| Stage II-IV vs metastatic | 0.62 [0.48; 0.75] | 0.53 [0.39; 0.67] |

### Multivariable analysis

The AUC was 0.89 (95%CI: 0.86-0.92) for model 1 (ADNEX predictors without CA125) and 0.92 (95%CI: 0.89-0.94) for model 5 (ADNEX predictors with CA125). Joint likelihood ratio tests for adding the cfDNA information to these models resulted in p-values of <0.001 (model 4 vs model 1) and 0.001 (model 8 vs model 5). The increase in AUC was 0.013 (95% CI 0.003 to 0.022) for model 4 vs 1, and 0.003 (-0.003 to 0.010) for model 8 vs 5 (**table 3**).

**Table 3.** Areas under the receiver operating curve (AUCs) with 95% confidence intervals for the multivariable models, and difference (delta) in AUC with 95% confidence intervals for models with cell-free DNA (cfDNA) information vs models without.

| **Model** | **Predictors** | **AUC** | **Delta AUC** |
| --- | --- | --- | --- |
| 1 | ADNEX without CA125 | 0.89 (0.86 to 0.92) | - |
| 2 | model 1 + gwz | 0.91 (0.88 to 0.93) | 0.014 (0.005 to 0.024) |
| 3 | model 1 + nucleosome | 0.90 (0.87 to 0.92) | 0.002 (-0.003 to 0.007) |
| 4 | model 1 + gwz + nucleosome | 0.91 (0.88 to 0.93) | 0.013 (0.003 to 0.022) |
| 5 | ADNEX with CA125 | 0.92 (0.89 to 0.94) | - |
| 6 | model 5 + gwz | 0.92 (0.89 to 0.94) | 0.005 (-0.002 to 0.012) |
| 7 | model 5 + nucleosome | 0.91 (0.89 to 0.94) | -0.002 (-0.004 to 0.001) |
| 8 | model 5 + gwz + nucleosome | 0.92 (0.89 to 0.94) | 0.003 (-0.003 to 0.01) |

Delta AUC: comparison of the AUC for models 2-4 vs model 1 and models 6-8 vs model 5.
gwz, genome-wide z-score.
ADNEX, Assessment of Different NEoplasias in the adneXa risk model

The increase in net benefit was small (**figure 2**, **supplementary tables 3-4**). At the 10% risk threshold, the increase was 0.0017 (95% CI: -0.0051 to 0.0086) for model 4 versus model 1, and 0.0020 (95% CI:

-0.0064 to 0.0098) for model 8 versus model 5. At this risk threshold, the test trade-off was 601 for model 4 versus model 1 and 494 for model 8 versus model 5. At this threshold, the lowest test tradeoff was 453 (model 2 vs model 1). Overall, the lowest test tradeoff was 71, when adding the genome-wide z-score to ADNEX variables without CA125 (model 2 vs model 1) at a risk threshold of 40%. In line with this, sensitivities and specificities for different risk thresholds were similar for models with and without cfDNA markers (**supplementary figure 5**, **supplementary table 5**). Depending on the risk threshold, sensitivity was between 0.014 lower (50% threshold) and 0.020 higher (40% threshold) for model 4 versus model 1, and specificity was between 0.004 (1% threshold) and 0.033 (50% threshold) higher.

For model 8 versus model 5, sensitivity was between -0.006 lower (20% threshold) and 0.009 higher (40% threshold), and specificity was between 0.014 lower (30% threshold) and 0.025 higher (40% threshold).

## DISCUSSION

In this study, the addition of two plasma cfDNA biomarkers to established clinical and ultrasound predictors from the ADNEX model added little to diagnostic performance. The univariable AUC values for the cfDNA markers were well below the reported AUC values of ultrasound-based prediction models such as ADNEX(3). While multivariable analysis yielded statistically significant likelihood ratio tests when adding the cfDNA markers to ADNEX variables, the increase in discrimination and clinical utility was marginal. When 10% is used as an acceptable threshold for the risk of malignancy to decide between specialized versus conservative surgery, adding cfDNA markers requires at least 453 patients for one extra true positive test result. Even when a threshold of 40% is considered acceptable, at least 71 patients for each true positive are required. Of note, an increase in test specificity using cfDNA-based biomarkers was not observed. We conclude that the cfDNA markers investigated in this study have no utility for improving the diagnosis of stage I invasive malignancies compared to the ADNEX model alone.

Strengths of our study are the sample size, the collection of clinical and ultrasound data according to a standardized protocol, the use of standardized procedures for plasma sampling and cfDNA extraction, and the control for intertest variability which is particularly important for cfDNA analyses relying on fragmentomics/nucleosomics(30). A first limitation is that the study was conducted in three oncology referral centers, resulting in high malignancy rates. For example, 16% of patients had borderline ovarian malignancy, which is a higher percentage than in previous IOTA studies (e.g. 7%(2)). It was previously observed that the diagnostic performance of cfDNA for borderline ovarian cancers is poor(13,14), possibly due to their low-grade, non-invasive biology usually linked to a lower stage and less extensive disease spread. A second limitation is that, despite the growing understanding of cfDNA biology, cfDNA testing often still suffers from a low signal-to-noise ratio(31). This might dilute the utility of cfDNA markers. The current study on cfDNA uses two previously developed biomarkers developed using genetic and non-genetic features from LC-WGS. Further improvements such as increasing test coverage might improve the current tests. At present, samples were sequenced with an average coverage of about 0.1x. An increase in sequencing coverage towards 1 to 1.5x (as performed in other plasma WGS studies(32)) might improve test performance. One can question the signal-to-noise ratio of the nucleosome score that we developed. We pooled genomic regions and assessed the average deviation of nucleosome patterns across the entire genome. We anticipate, however, that focusing the score on genomic regions specifically altered in HGSOC or non-HGSOC could still improve the performance. A third limitation is that, despite the use of standardized procedures, cfDNA extraction was carried out by two different laboratories. This may have further affected the signal-to-noise ratio by introducing additional variability.

Two other considerations are worth mentioning. First, to calculate the cfDNA markers, one needs the reference samples from healthy individuals and patients with ovarian cancer. It remains to be investigated how different sets of reference samples will affect the scores and their performances. The nucleosome score in our study was derived using cfDNA data from patients with high-grade serous ovarian cancer as a positive control. Using different control sets may lead to different scores, which may lead to different risk estimates. Secondly, the patients in this study were scheduled for surgery. Hence, the results only assess the added value of cfDNA in patients where the decision to operate was already taken.

Our findings have implications for the implementation of cfDNA into the early diagnosis of ovarian cancer. We focused on the genome-wide z-score(13), which leverages genetic features such as the presence of cancer-derived CNAs in cfDNA (‘second generation’ type of liquid biopsy(33)), and the nucleosome score(14), which focuses on non-genetic feature such as the quantification of tumor-specific cfDNA fragmentation (‘third generation’ type of liquid biopsy(33)). If clinical utility is currently lacking in high-risk patients (i.e. patients with adnexal masses), one could argue whether cfDNA analysis could be used in the future in a general population for early cancer detection. However, these findings should encourage the field to improve the clinical utility of cfDNA-based detection. Current advances in cfDNA research also focus on the interrogation of a larger number of features obtained from plasma WGS data using machine learning algorithms (cf. ‘fourth generation’ liquid biopsies(33)). A recent study included machine learning on plasma WGS for the detection of mutational processes and reported an AUC of 0.96 to distinguish patients with stage I-IV colorectal cancer from healthy controls(32). Importantly, developing and evaluating these cfDNA-based biomarkers heavily depends on using well-described training and testing datasets with clearly-defined inclusion criteria beyond tumor stage. The dataset in our study allows for further development of these biomarkers.

Our study indicates that cfDNA analysis on pre-operative plasma samples, using two read-outs focused on the aggregate detection of chromosomal instability (genome-wide z-score) and nucleosome footprinting of cfDNA (nucleosome score), has limited clinical utility in addition to established clinical and ultrasound-based predictors for discriminating between benign and malignant ovarian masses.

## DATA AVAILABILITY

The pseudonymized data and data dictionary are stored in the KU Leuven Research Data Repository (RDR), https://doi.org/10.48804/TXL95Z. The dataset is not publicly available because this was not part of the informed consent. However, the dataset may be obtained following permission of prof. AC and prof. DT and after fulfilling all data transfer requirements.

## ACKNOWLEDGEMENTS

The authors would like to thank Annick Van den Broeck, Gitte Thirion and Katja Vandenbrande for technical and logistic support.

