## Supplementary material for "Added value of cell-free DNA over clinical and ultrasound information for diagnosing ovarian cancer"

### SUPPLEMENTARY APPENDIX 1: DETAILS ON IMPUTATION AND STATISTICAL ANALYSIS

#### 1. IMPUTATION OF MISSING VALUES

We used single stochastic imputation based on 'multivariate imputation by chained equations' to address the missing values of genome-wide z-score, nucleosome score and CA125(1). The imputation was done using predictive mean matching regression. Variables in the imputation model are: patient age (in years), maximum diameter of the lesion (in mm)(log-transformed), proportion solid tissue (calculated as the maximum diameter of the largest solid component in mm, divided by the maximum diameter of the lesion in mm) (with a linear and a quadratic term), number of papillations (ordinal variable: 0, 1, 2, 3, >3), more than 10 locules (yes/no), presence of ascites (yes/no), presence of acoustic shadows (yes/no), CA125 (log-log transformed, only used to impute genome-wide z-score and nucleosome score), genome-wide z-score (Box-Cox transformation with lambda -0.3, only used to impute CA125 and nucleosome score), nucleosome score (only used to impute CA125 and genome-wide z-score), the protocol (cf. lab) used for the DNA analysis and outcome (benign, borderline, stage I primary invasive, stage II-IV primary invasive and metastatic).

**Figure 1.1. Convergence plots for CA125, genome-wide z-score and nucleosome score.** The plots on the top row refer to nucleosome score, the plots in the middle refer to CA125 (after log-log transformation; T_CA125) and the plot on the bottom row refer to genome-wide z-score (after boxcox-transformation with lambda -0.3; T_GWZ). The plots on the left show the mean value, the plots on the right show the standard deviation (sd). The x-axis refers to the iteration (1 to 100).

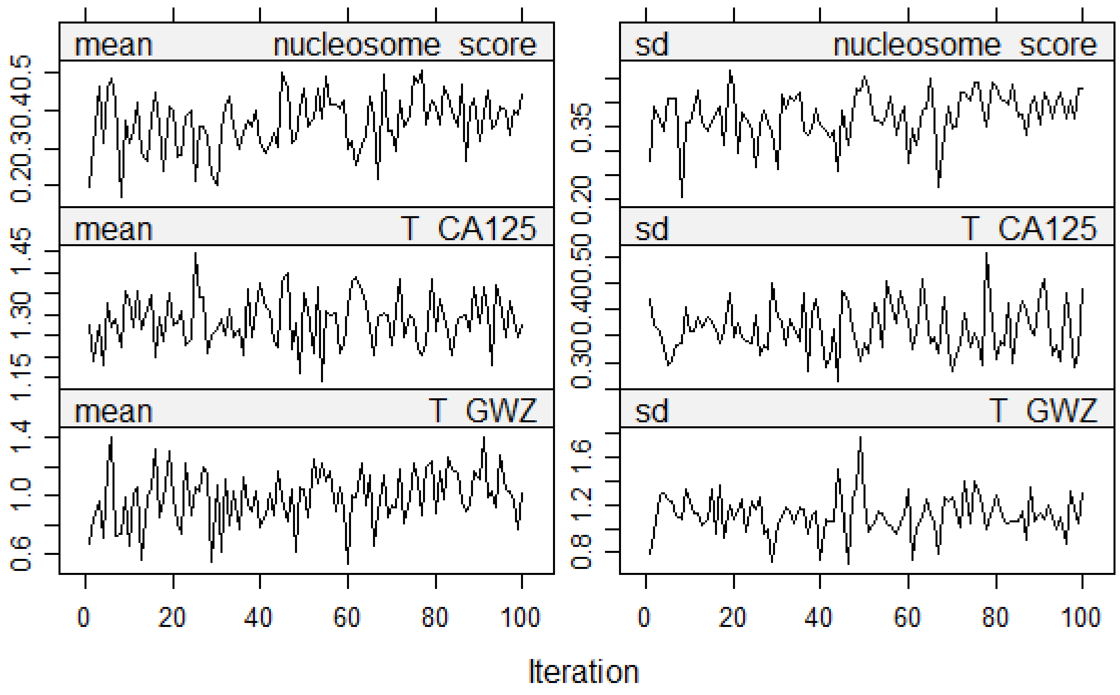

**Figure 1.2. Density plots of nucleosome score, log(log(CA125+1)) and (genome-wide z-score$^{-0.3}$ – 1)/-0.3.** The blue curve is the density of the values that were observed (i.e. were not missing), the red curves are the density for the imputation of the values that were missing.

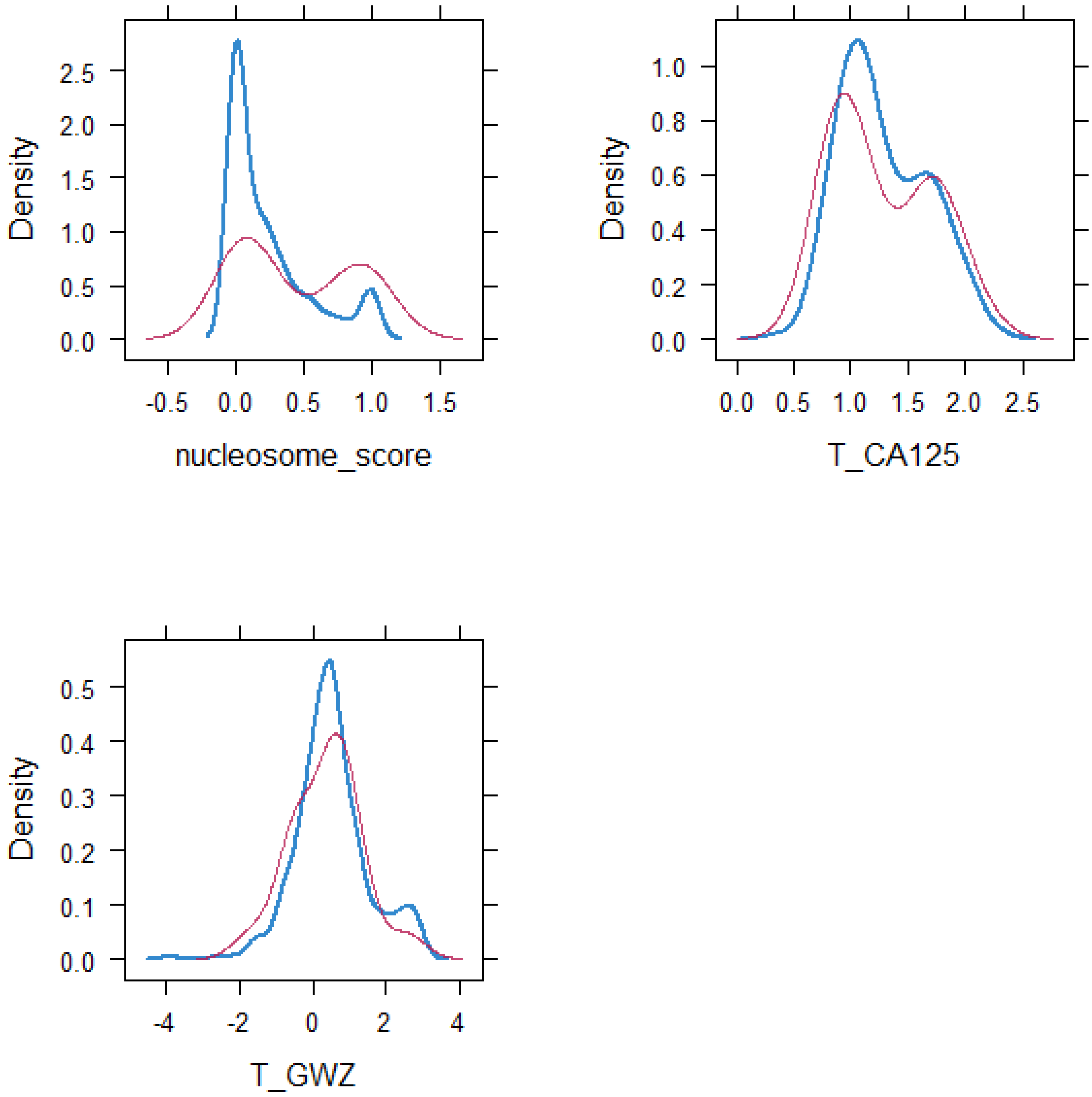

### 2. DETAILS ABOUT THE LOGISTIC MODELS

Logistic regression models using Firth's correction were fitted to look at the added diagnostic value of the cfDNA scores in combination with established clinical and ultrasound predictors from the ADNEX model(2). The predictor 'type of center' was not used, because the three participating centers were of the same type (i.e. oncological center). The eight predictors were modeled exactly as in the ADNEX model: age (no transformation), maximum diameter of lesion ($log_2$-transformed), proportion of solid tissue and its quadratic term, presence of more than 10 cyst locules (binary), number of papillary projections (linear instead of categorical), presence of acoustic shadows (binary), presence of ascites (binary) and CA125 ($log_2$-transformed).

### 3. SAMPLE SIZE DETERMINATION

The available sample size is 526, which is more or less evenly split between patients with a benign (272) vs malignant (254) tumor. A sample size calculation for the power of the joint likelihood ratio test is not straightforward. We conducted a simple simulation study with following assumptions:

- We simulated 1 variable to represent the linear predictor (LP) for the ADNEX variables, and 2 variables to represent the z and nucleosome scores
- All variables are simulated as having a normal distribution in patients with a benign vs malignant tumor. In particular, all variables are simulated as having a N(0,1) distribution in patients with a benign tumor, and as having a N(mu,1) distribution in patients with a malignant tumor
- The correlation between the z and nucleosome scores is assumed to be 0.6 (both in patients with a benign tumor and in patients with a malignant tumor). This correlation is based on results presented in Vanderstichele et al (2022)(3)
- The alpha level is 0.05, the sample size is 526 (272 benign, 254 malignant)

We have to specify mu for the ADNEX LP (mu1), z score (mu2), and nucleosome score (mu3), and correlations between the ADNEX LP and the cfDNA variables. We choose mu1 to be either 1.81 or 2.087. This corresponds to AUCs of 0.90 and 0.93, respectively, for the ADNEX variables. These seem reasonable settings based on existing external validation studies of ADNEX (e.g. Van Calster et al, BMJ 2020(4)). We set mu2 = mu3, and vary between values of 0.358, 0.74, and 1.19. These three values correspond to AUCs of 0.60, 0.70 and 0.80, respectively. Vanderstichele et al (2022)(3) report AUCs of around 0.7, so this is reasonable. The correlation between the ADNEX LP and the cfDNA variables is unclear. We use values of 0.2, 0.5, and 0.8, to cover a wide range. Combining the choices for mu1, mu2/mu3, and the correlation leads to 18 scenarios. For each scenario, we simulate 2000 datasets, and report the proportion of datasets for which the joint likelihood ratio test was significant. This is an estimate of the power. In addition, we simulate for each scenario a large dataset consisting of 2 million patients (50% benign, 50% malignant), fit models with only the ADNEX LP or with all three variables, and evaluate the AUC. These AUCs should reflect the true AUCs given the large dataset.

For the models that only include the ADNEX LP, the AUCs should be either 0.90 or 0.93 depending on the choice for mu1. This allows us to report the true increase in AUC following the addition of the cfDNA variables. The results of the simulations are presented in Table 1.1.

Table 1.1. Results for the simulation study to investigate power for the joint likelihood ratio test.

| **Mu1** | **Mu2/mu3** | **Correlation** | **Power** | **True AUC increase** |
|---|---|---|---|---|
| 1.81 | 0.358 | 0.2 | 5% | <0.001 |
| 1.81 | 0.358 | 0.5 | >99% | 0.017 |
| 1.81 | 0.358 | 0.8 | >99% | 0.090 |
| 1.81 | 0.74 | 0.2 | 82% | 0.006 |
| 1.81 | 0.74 | 0.5 | 28% | 0.002 |
| 1.81 | 0.74 | 0.8 | >99% | 0.063 |
| 1.81 | 1.19 | 0.2 | >99% | 0.026 |
| 1.81 | 1.19 | 0.5 | 74% | 0.005 |
| 1.81 | 1.19 | 0.8 | >99% | 0.013 |
| 2.087 | 0.358 | 0.2 | 6% | <0.001 |
| 2.087 | 0.358 | 0.5 | >99% | 0.017 |
| 2.087 | 0.358 | 0.8 | >99% | 0.067 |
| 2.087 | 0.74 | 0.2 | 58% | 0.003 |
| 2.087 | 0.74 | 0.5 | 67% | 0.004 |
| 2.087 | 0.74 | 0.8 | >99% | 0.056 |
| 2.087 | 1.19 | 0.2 | >99% | 0.016 |
| 2.087 | 1.19 | 0.5 | 19% | 0.001 |
| 2.087 | 1.19 | 0.8 | >99% | 0.026 |

The results indicate that the correlation between the ADNEX LP and the cfDNA variables is crucial, and has perhaps an unexpected impact. For high correlations, there is more power when the univariable effects of the cfDNA variables are smaller. Importantly, however, the results suggest that power reaches about 80% when the true AUC increase is 0.006. This suggests that the sample size is sufficiently informative for the first examination of added value of cfDNA. This sample size investigation is not perfect, because the assumption of normally distributed variables in patients with benign vs malignant tumors does not hold in reality.

If we apply the sample size calculation procedure for prediction model development studies as described in Riley et al (2020)(5), we obtain a minimum sample size of 384. This was obtained using the pmsampsize package in R, using the following specifications: 13 parameters (9 for the ADNEX variables including CA125, 4 for the cfDNA parameters including spline modeling), 48% event fraction, and a Cox-Snell R-squared of 0.444. This value for the R-squared approximates an AUC of 0.9 (Riley,

2021(6)). Although the aim of this study is not to develop a prediction model, this sample size analysis further supports the available sample size for studying the added value of the cfDNA scores.

**Supplementary Table 1 | cfDNA score for the included patients.** Genome-wide z-score and nucleosome scores according to histological outcome categories.

| Variable | Median (IQR) and range |
|---|---|
| Observed genome wide z-score | |
| Overall | 0.662 (-0.023 – 2.354) |
| | Range: -0.925 – 4030.589 |
| Benign | 0.29 (IQR -0.22 – 0.90) |
| | Range: -0.93 – 93.11 |
| Borderline | 0.54 (IQR 0.02 – 1.42) |
| | Range: -0.86 – 135.18 |
| Stage I invasive | 0.89 (IQR 0.19 – 1.78) |
| | Range: -0.71 – 458.58 |
| Stage II-IV invasive | 7.57 (IQR 1.40 – 47.00) |
| | Range: -0.73 – 4030.59 |
| Metastatic | 0.81 (IQR 0.10 – 232.11) |
| | Range: -0.62 – 765.58 |
| Observed nucleosome score | |
| Overall | 0.1081 (IQR 0.0019 - 0.3606) |
| | Range: 0.0002 - 1 |
| Benign | 0.0546 (IQR 0.0013 - 0.2387) |
| | Range: 0.0002 - 0.9995 |
| Borderline | 0.0556 (IQR 0.0024 - 0.2970) |
| | Range: 0.0004 - 0.9997 |
| Stage I invasive | 0.0945 (IQR 0.0024 - 0.2461) |
| | Range: 0.0004 - 0.9999 |
| Stage II-IV invasive | 0.4393 (IQR 0.1029 - 0.9322) |
| | Range: 0.0004 - 1 |
| Metastatic | 0.2450 (IQR 0.0016 - 0.9930) |
| | Range: 0.0003 - 0.9999 |

IQR, interquartile range

**Supplementary Table 2 | Correlation between ADNEX-variables and the cfDNA scores.** The correlations with age, maximum diameter of lesion, proportion of solid tissue, number of papillary projections and CA125 is calculated with the Spearman correlation. The correlations with presence of more than 10 locules, presence of acoustic shadows and presence of ascites is calculated with the point biserial correlation.

| | Genome-wide z-score | Nucleosome score |
|---|---|---|
| Age | 0.21 | 0.15 |
| Maximum Diameter Lesion | 0.02 | 0.03 |
| Proportion solid tissue | 0.30 | 0.16 |
| > 10 locules | 0.01 | -0.01 |
| Number papillary projections | -0.05 | -0.02 |
| Acoustic shadows | 0.09 | 0.13 |
| Ascites | -0.16 | -0.35 |
| CA125 | 0.33 | 0.28 |

**Supplementary Table 3 | Net benefit of models 1-4 (without CA125) for the comparison benign (n=272) versus malignant (n=254).**

| Risk threshold | Model | NB | Delta NB vs model 1 (95% CI) | Test tradeoff |
|---|---|---|---|---|
| 5% | Model 1 | 0.459 | | |
| | Model 2 | 0.460 | 0.0002 (-0.0006 to 0.0036) | 4226 |
| | Model 3 | 0.459 | -0.0002 (-0.0032 to 0.0018) | Infinite |
| | Model 4 | 0.460 | 0.0001 (-0.0012 to 0.0037) | 8105 |
| 10% | Model 1 | 0.438 | | |
| | Model 2 | 0.441 | 0.0022 (-0.0043 to 0.0088) | 453 |
| | Model 3 | 0.439 | 0.0012 (-0.0032 to 0.0067) | 818 |
| | Model 4 | 0.441 | 0.0017 (-0.0051 to 0.0086) | 601 |
| 20% | Model 1 | 0.402 | | |
| | Model 2 | 0.402 | -0.0002 (-0.0093 to 0.0159) | Infinite |
| | Model 3 | 0.404 | 0.0017 (-0.0069 to 0.0145) | 589 |
| | Model 4 | 0.401 | -0.001 (-0.0096 to 0.0175) | Infinite |
| 30% | Model 1 | 0.365 | | |
| | Model 2 | 0.371 | 0.0073 (-0.0082 to 0.0241) | 138 |
| | Model 3 | 0.361 | -0.0036 (-0.0101 to 0.0176) | Infinite |
| | Model 4 | 0.370 | 0.0062 (-0.0095 to 0.0231) | 161 |
| 40% | Model 1 | 0.334 | | |
| | Model 2 | 0.349 | 0.0140 (-0.0081 to 0.0324) | 71 |
| | Model 3 | 0.337 | 0.0021 (-0.0156 to 0.0161) | 467 |
| | Model 4 | 0.347 | 0.0120 (-0.0095 to 0.0311) | 83 |

NB, net benefit.

**Supplementary Table 4 | Net benefit of models 5-8 (with CA125) for the comparison benign (n=272) versus malignant (n=254).**

| Risk threshold for ovarian malignancy | Model | NB | Delta NB vs model 5 (95% CI) | Test tradeoff |
|---|---|---|---|---|
| 5% | Model 5 | 0.456 | | |
| | Model 6 | 0.459 | 0.0022 (-0.0025 to 0.006) | 450 |
| | Model 7 | 0.458 | 0.0015 (-0.0023 to 0.0051) | 658 |
| | Model 8 | 0.457 | 0.0016 (-0.0039 to 0.0061) | 636 |
| 10% | Model 5 | 0.438 | | |
| | Model 6 | 0.441 | 0.0004 (-0.0053 to 0.0101) | 2489 |
| | Model 7 | 0.439 | -0.0026 (-0.0054 to 0.006) | Infinite |
| | Model 8 | 0.441 | 0.002 (-0.0064 to 0.0098) | 494 |
| 20% | Model 5 | 0.402 | | |
| | Model 6 | 0.402 | -0.0017 (-0.0069 to 0.015) | Infinite |
| | Model 7 | 0.404 | -0.0012 (-0.0074 to 0.0097) | Infinite |
| | Model 8 | 0.401 | -0.0023 (-0.008 to 0.0149) | Infinite |
| 30% | Model 5 | 0.365 | | |
| | Model 6 | 0.371 | 0.0027 (-0.0095 to 0.0190) | 370 |
| | Model 7 | 0.361 | -0.0014 (-0.0095 to 0.0103) | Infinite |
| | Model 8 | 0.370 | 0.0002 (-0.0104 to 0.0208) | 6294 |
| 40% | Model 5 | 0.334 | | |
| | Model 6 | 0.349 | 0.0117 (-0.0079 to 0.0269) | 85 |
| | Model 7 | 0.337 | 0.0016 (-0.0117 to 0.0124) | 614 |
| | Model 8 | 0.347 | 0.0129 (-0.0087 to 0.0281) | 78 |

NB, net benefit.

95% CI, 95% confidence interval

**Supplementary Table 5 | Sensitivity and specificity of the different models for the comparison benign (n=272) versus malignant (n=254).** Sensitivities and specificities according to different risk thresholds for ovarian malignancy.

| Risk threshold for ovarian malignancy | Model | Sensitivity (95% CI) | Specificity (95% CI) |
|---|---|---|---|
| 1% | Model 1 | 99.9 (83.7 to 100) | 8.1 (5.3 to 11.9) |
| | Model 2 | 99.9 (83.7 to 100) | 9.1 (6.2 to 13.2) |
| | Model 3 | 99.9 (83.7 to 100) | 6.2 (3.9 to 9.8) |
| | Model 4 | 99.9 (83.7 to 100) | 8.5 (5.7 to 12.4) |
| | Model 5 | 99.9 (83.7 to 100) | 4.8 (2.8 to 8.0) |
| | Model 6 | 99.9 (83.7 to 100) | 5.5 (3.3 to 8.9) |
| | Model 7 | 99.9 (83.7 to 100) | 4.4 (2.5 to 7.6) |
| | Model 8 | 99.9 (83.7 to 100) | 6.6 (4.1 to 10.2) |
| 5% | Model 1 | 99.6 (97.2 to 99.9) | 27.8 (22.8 to 33.5) |
| | Model 2 | 99.6 (97.2 to 99.9) | 28.5 (23.5 to 34.2) |
| | Model 3 | 99.5 (97.2 to 99.9) | 27.5 (22.5 to 33.1) |
| | Model 4 | 99.6 (97.2 to 99.9) | 28.5 (23.5 to 34.2) |
| | Model 5 | 98.7 (96.2 to 99.5) | 30.7 (25.5 to 36.5) |
| | Model 6 | 99.0 (96.7 to 99.6) | 32.6 (27.3 to 38.4) |
| | Model 7 | 99.0 (96.7 to 99.6) | 30.0 (24.8 to 35.7) |
| | Model 8 | 99.0 (96.6 to 99.5) | 31.4 (26.2 to 37.2) |
| 10% | Model 1 | 98.2 (95.7 to 99.2) | 40.2 (34.5 to 46.1) |
| | Model 2 | 98.5 (96.1 to 99.3) | 41.6 (35.9 to 47.6) |
| | Model 3 | 98.6 (96.2 to 99.4) | 39.1 (33.4 to 45.0) |
| | Model 4 | 98.5 (96.0 to 99.3) | 41.2 (35.5 to 47.2) |
| | Model 5 | 97.2 (94.4 to 98.5) | 43.4 (37.6 to 49.4) |
| | Model 6 | 97.2 (94.4 to 98.5) | 44.6 (38.8 to 50.5) |
| | Model 7 | 96.8 (93.8 to 98.2) | 42.7 (36.9 to 48.7) |
| | Model 8 | 97.5 (94.8 to 98.7) | 44.5 (38.6 to 50.4) |
| 20% | Model 1 | 95.1 (91.8 to 97.1) | 56.8 (50.9 to 62.6) |
| | Model 2 | 94.3 (90.8 to 96.4) | 59.8 (53.9 to 65.4) |
| | Model 3 | 95.0 (91.7 to 96.9) | 58.6 (52.6 to 64.3) |
| | Model 4 | 94.2 (90.7 to 96.3) | 59.7 (53.7 to 65.3) |
| | Model 5 | 94.4 (90.9 to 96.5) | 62.4 (56.5 to 67.9) |
| | Model 6 | 93.9 (90.3 to 96.1) | 63.0 (57.1 to 68.5) |
| | Model 7 | 94.2 (90.8 to 96.4) | 61.9 (56.0 to 67.5) |
| | Model 8 | 93.8 (90.2 to 96.0) | 62.9 (57.0 to 68.4) |
| 30% | Model 1 | 91.5 (87.4 to 94.2) | 65.9 (60.0 to 71.2) |
| | Model 2 | 91.8 (87.9 to 94.5) | 68.3 (62.6 to 73.5) |
| | Model 3 | 90.5 (86.3 to 93.3) | 66.5 (60.7 to 71.8) |
| | Model 4 | 91.7 (87.7 to 94.3) | 68.2 (62.4 to 73.4) |
| | Model 5 | 90.6 (86.5 to 93.5) | 74.3 (68.8 to 79.1) |
| | Model 6 | 91.4 (87.4 to 94.1) | 73.8 (68.3 to 78.6) |
| | Model 7 | 90.1 (85.9 to 93.0) | 74.9 (69.4 to 79.6) |
| | Model 8 | 91.3 (87.3 to 94.0) | 72.9 (67.3 to 77.8) |
| 40% | Model 1 | 85.8 (81.1 to 89.5) | 77.4 (72.1 to 81.9) |
| | Model 2 | 88.1 (83.6 to 91.4) | 78.4 (73.2 to 82.8) |
| | Model 3 | 86.4 (81.7 to 89.9) | 77.3 (72.0 to 81.8) |
| | Model 4 | 87.8 (83.3 to 91.1) | 78.1 (72.9 to 82.5) |
| | Model 5 | 85.5 (80.7 to 89.2) | 79.7 (74.5 to 84.0) |
| | Model 6 | 86.5 (81.9 to 90.1) | 81.7 (76.7 to 85.7) |
| | Model 7 | 85.7 (81.0 to 89.4) | 79.8 (74.7 to 84.1) |
| | Model 8 | 86.4 (81.7 to 89.9) | 82.2 (77.3 to 86.2) |
| 50% | Model 1 | 81.0 (75.7 to 85.2) | 82.6 (77.7 to 86.5) |
| | Model 2 | 80.0 (74.7 to 84.4) | 86.9 (82.4 to 90.3) |
| | Model 3 | 79.6 (74.3 to 84.0) | 83.1 (78.3 to 87.0) |
| | Model 4 | 79.7 (74.5 to 84.1) | 85.9 (81.4 to 89.4) |
| | Model 5 | 83.6 (78.6 to 87.6) | 84.3 (79.6 to 88.1) |
| | Model 6 | 84.8 (80.0 to 88.6) | 87.3 (82.9 to 90.6) |
| | Model 7 | 82.3 (77.2 to 86.4) | 85.6 (81.0 to 89.1) |
| | Model 8 | 84.3 (79.4 to 88.1) | 86.6 (82.2 to 90.0) |

**Supplementary Figure 1 | Boxplot visualization of the distribution of the genome-wide z-score**

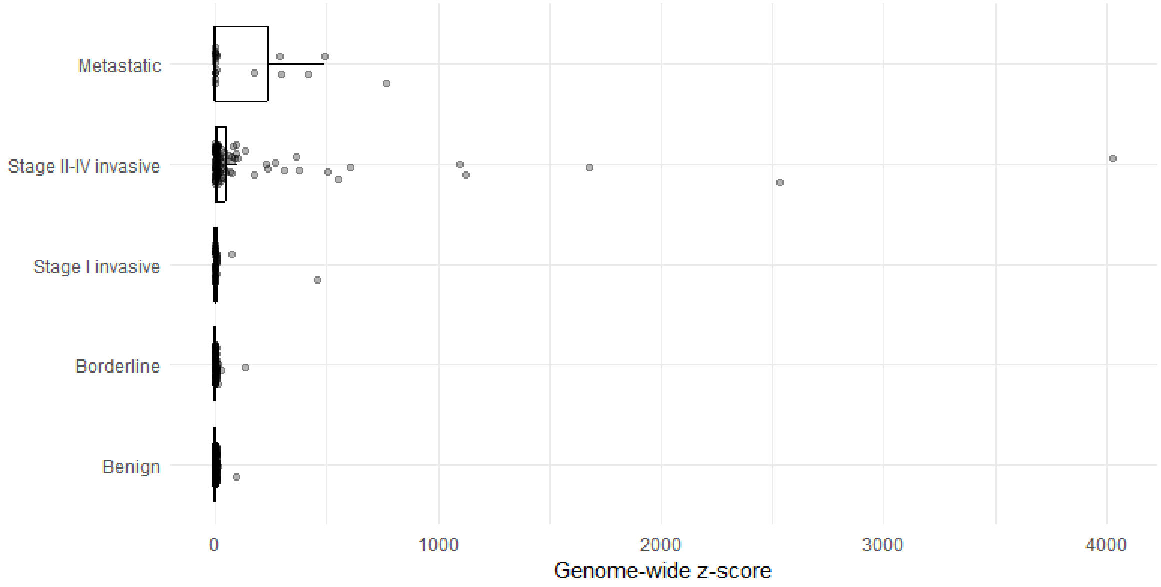

**Supplementary Figure 2 | Boxplot visualization of the distribution of the nucleosome score**

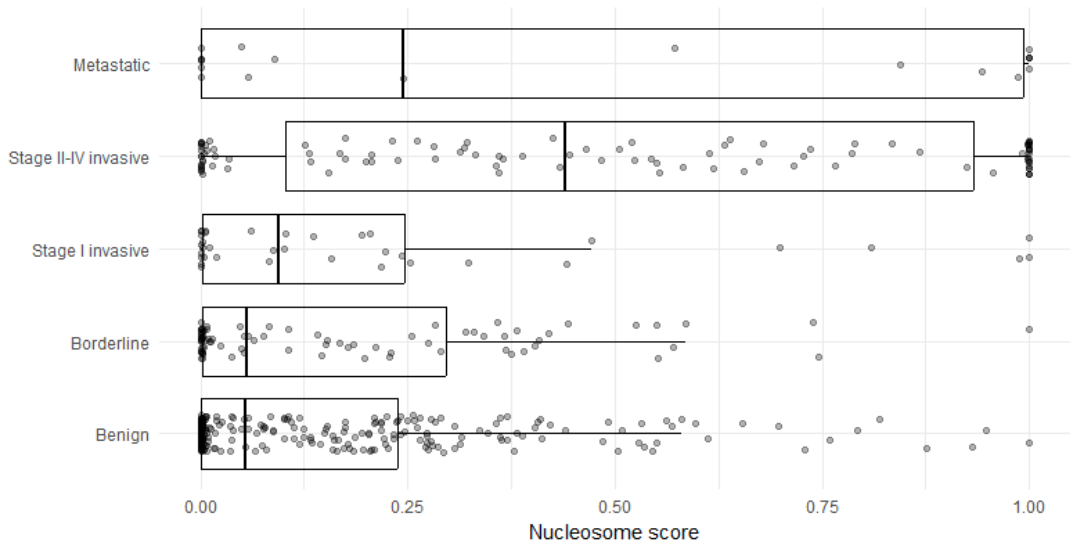

**Supplementary Figure 3 | Bland-Altman plot showing the difference in the genome-wide z-score between the two protocols.** The confidence interval limits for the mean difference is shown in blue and for the limits of agreement in green and red.

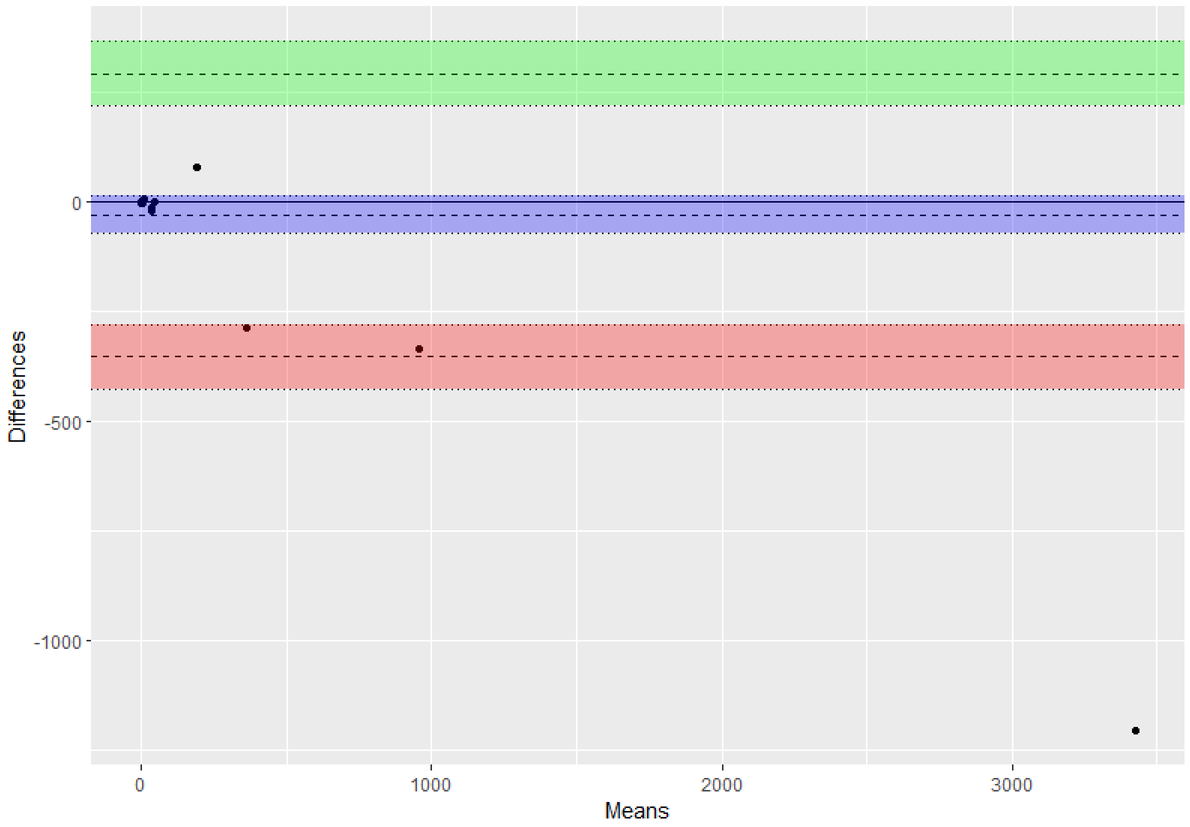

**Supplementary Figure 4 | Bland-Altman plot showing the difference in the nucleosome score between the two protocols.** The confidence interval limits for the mean difference is shown in blue and for the limits of agreement in green and red.

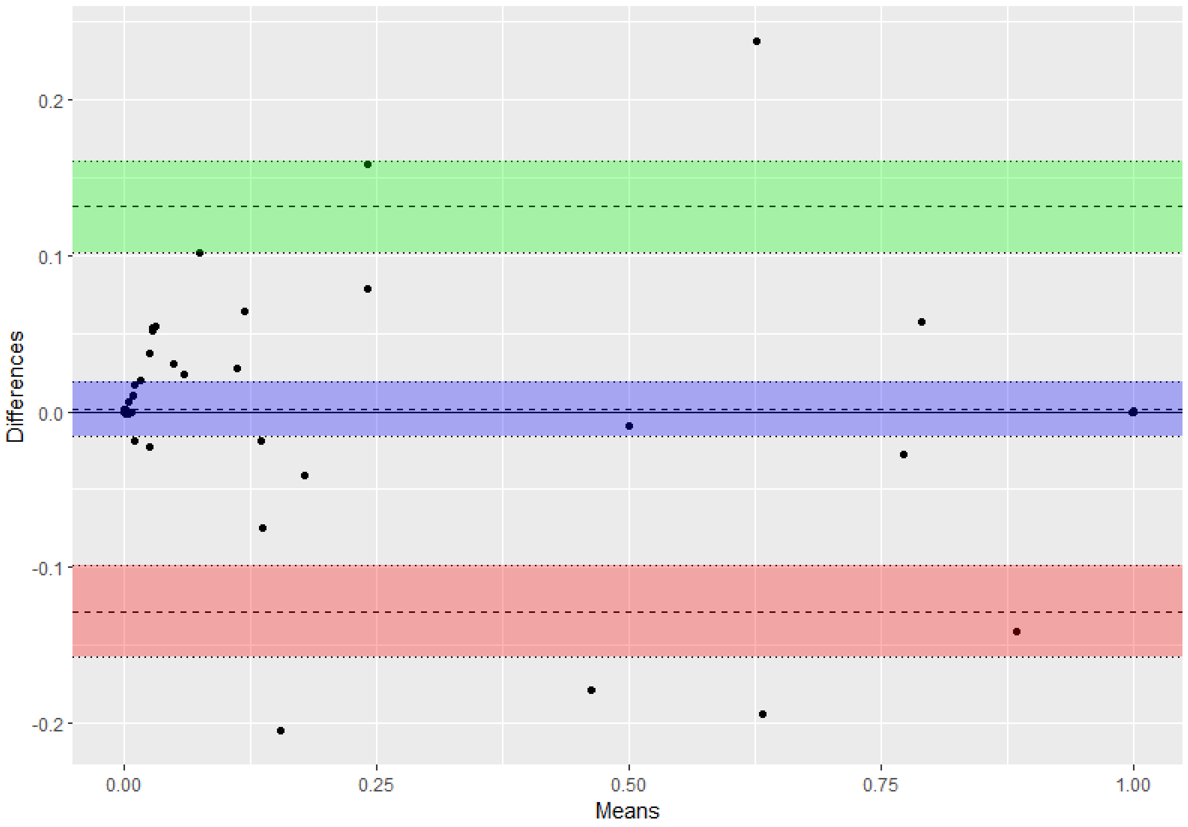

**Supplementary Figure 5 | Classification plot showing sensitivity and specificity for models 1-8.**

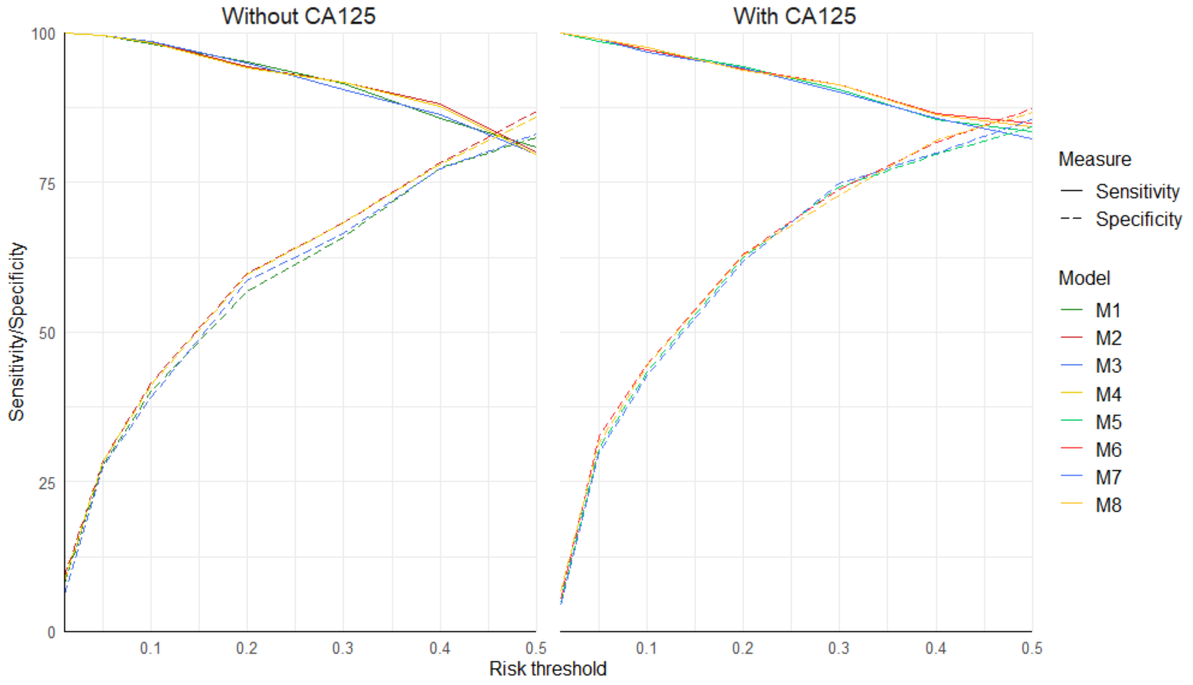
